# The Athens-Luebeck Questionnaire: a tool to discriminate between subtypes of Persistent Postural Perceptual Dizziness

**DOI:** 10.1101/2025.01.30.25321410

**Authors:** Evangelos Anagnostou, Georgios Armenis, Athena Zachou, Renana Storm, Andreas Sprenger, Christoph Helmchen

## Abstract

**Introduction:** Persistent Postural-Perceptual Dizziness (PPPD), as an umbrella term for functional dizziness, encompasses a wide range of subjective symptoms affecting visual, vestibular, and motor functions. We developed the Athens-Lübeck Questionnaire (ALQ) as a bedside tool to differentiate specific symptom subtypes, which could inform more targeted research into the pathogenesis of the syndrome and facilitate tailored physiotherapeutic interventions.

**Methods:** A total of 96 patients with primary or secondary PPPD were included in a prospective cross-sectional study conducted at two tertiary referral centers. All participants had unimpaired vestibular function, as verified by video head-impulse testing at the time of examination. Each participant completed the ALQ, an 8-item questionnaire divided into four symptom subtypes: ALQvis (visual intolerance), ALQstand (intolerance to quiet standing or sitting), ALQpass (passive motion intolerance), and ALQact (active motion intolerance). We assessed the reliability of the questionnaire, the prevalence of different symptom subtypes, and the presence of dominant symptom profiles.

**Results:** The ALQ demonstrated good internal consistency, with a Cronbach’s alpha of 0.813. Items within the same symptom domain showed strong inter-item correlations. Approximately two-thirds of the participants exhibited a predominant symptom subtype, with the majority classified under the ALQact phenotype.

**Conclusion:** The 8-item ALQ is a valid tool for identifying distinct PPPD symptom subtypes. Its primary strengths lie in its brevity and ease of use in outpatient vertigo clinics, enabling the identification of predominant phenotypes that may be relevant for guiding tailored therapeutic interventions.

## Introduction

The definition of Persistent Postural-Perceptual Dizziness (PPPD), established through a seminal consensus by numerous balance specialists in 2017, marked a new era in diagnosing and treating patients with functional dizziness (1). By integrating various preexisting conceptualizations of non-organic dizziness—such as phobic postural vertigo, space motion discomfort, visual vertigo, and chronic subjective dizziness (2–8)—PPPD redefined functional dizziness and moved away from older psychiatric connotations like psychogenic and psychosomatic dizziness. These older terms do not acknowledge the various physiological and behavioral abnormalities evident in PPPD (9–19) and overlook that a substantial portion of PPPD patients do not suffer from depression or anxiety disorders.

However, clinicians who see many PPPD sufferers often feel that there are substantial differences from patient to patient under the umbrella of functional dizziness. These differences may hinder the application of a simplistic, one-size-fits-all treatment approach, which may neglect the dominance of certain symptoms over others. Therefore, it might be significant to consider whether patients developed primary PPPD or secondary PPPD (e.g. due to an initial organic vestibular disorder), or whether there is a comorbid anxiety disorder. Moreover, the predominance of specific symptoms of functional dizziness is crucial. In our experience, some patients primarily complain of dizziness when passively moved, for example, in a vehicle, but not when standing still or actively moving while walking. Others experience discomfort and imbalance when standing upright, which improves when they start moving. Still, others report that their principal dizziness and disorientation symptoms emerge when exposed to large moving visual stimuli.

Applying a rigid pharmaceutical or physiotherapeutic treatment regimen to such different phenotypes may miss the opportunity to address the patient’s dominant complaint, thus reducing therapeutic success. Currently, there are no clinical tools to separate such PPPD subtypes, although the Niigata PPPD Questionnaire (NPQ) can be considered an indirect attempt to address the presence of subtypes (20, 21). However, the NPQ was designed to assist in diagnosing PPPD and assessing its severity rather than directly identifying PPPD subtypes (20).

Here, we aimed to directly address four main symptom subtypes of PPPD, namely: the phenotype characterized by visual intolerance; the subtype primarily complaining of dizziness and imbalance while sitting or standing still upright, often with improvement during self-motion or physical exercise; the subtype whose symptoms worsen with passive movement; and the so-called active type, who experiences increased dizziness during active movement. To this end, we developed the Athens-Lübeck Questionnaire (ALQ), which contains four domains with direct questions regarding the putative four subtypes.

To better capture each patient’s idiosyncrasies and level of comprehension and thereby avoid misunderstandings, we constructed two conceptually similar questions for each domain. These questions address the same symptoms with slightly altered phrasing to reduce the possibility of misinterpretation. We then assessed the internal consistency of the questionnaire before testing for interrelations among the different items. Finally, we proceeded to the main goal of this study: characterizing the frequency of the different symptom subtypes as identified by the ALQ and determining whether subtypes with physiologically distinct phenomenology exist.

## Methods

A prospective study was conducted on patients diagnosed with PPPD based on the 2017 Bárány Society criteria (1). Participants were referred to either the Outpatient Clinic for Vertigo and Balance Disorders at the University of Athens or the Vertigo Clinic at the University of Lübeck.

The study was conducted in accordance with the Declaration of Helsinki and received approval from the Ethics Committees of the Neurology Departments at both the University of Athens and the University of Lübeck (17-036, AZ 21-098).

All subjects underwent a comprehensive neurological, neuro-otological, and neuro-ophthalmological clinical examination. Each patient also underwent video head impulse testing (vHIT) and a brain MRI. Any abnormalities detected in these clinical or instrumental assessments were considered exclusion criteria for the study. Additionally, several patients underwent caloric labyrinthine testing, posturography, audiogram testing, subjective visual vertical testing, or vestibular evoked myogenic potential (VEMP) testing.

All patients were asked to complete the Athens-Lübeck Questionnaire (ALQ, Table 1), which was developed to test the hypothesis that specific PPPD phenotypes or symptom subtypes predominate over others. The ALQ consists of four domains, each containing direct questions targeting the four putative symptom subtypes. Two conceptually similar questions were included for each subtype to ensure the intended meaning was clearly conveyed to the patient and to address different facets of understanding, thereby reducing the likelihood of misinterpretation. Each question represented one questionnaire item, resulting in two items per symptom subtype. A 6-point Likert scale was used for each item, ranging from “never” to “unbearable” in severity. The four symptom subtypes are as follows:

- ALQvis: Visual intolerance
- ALQstand: Intolerance to quiet standing or sitting
- ALQpass: Passive motion intolerance
- ALQact: Active motion intolerance

**Table 1.**
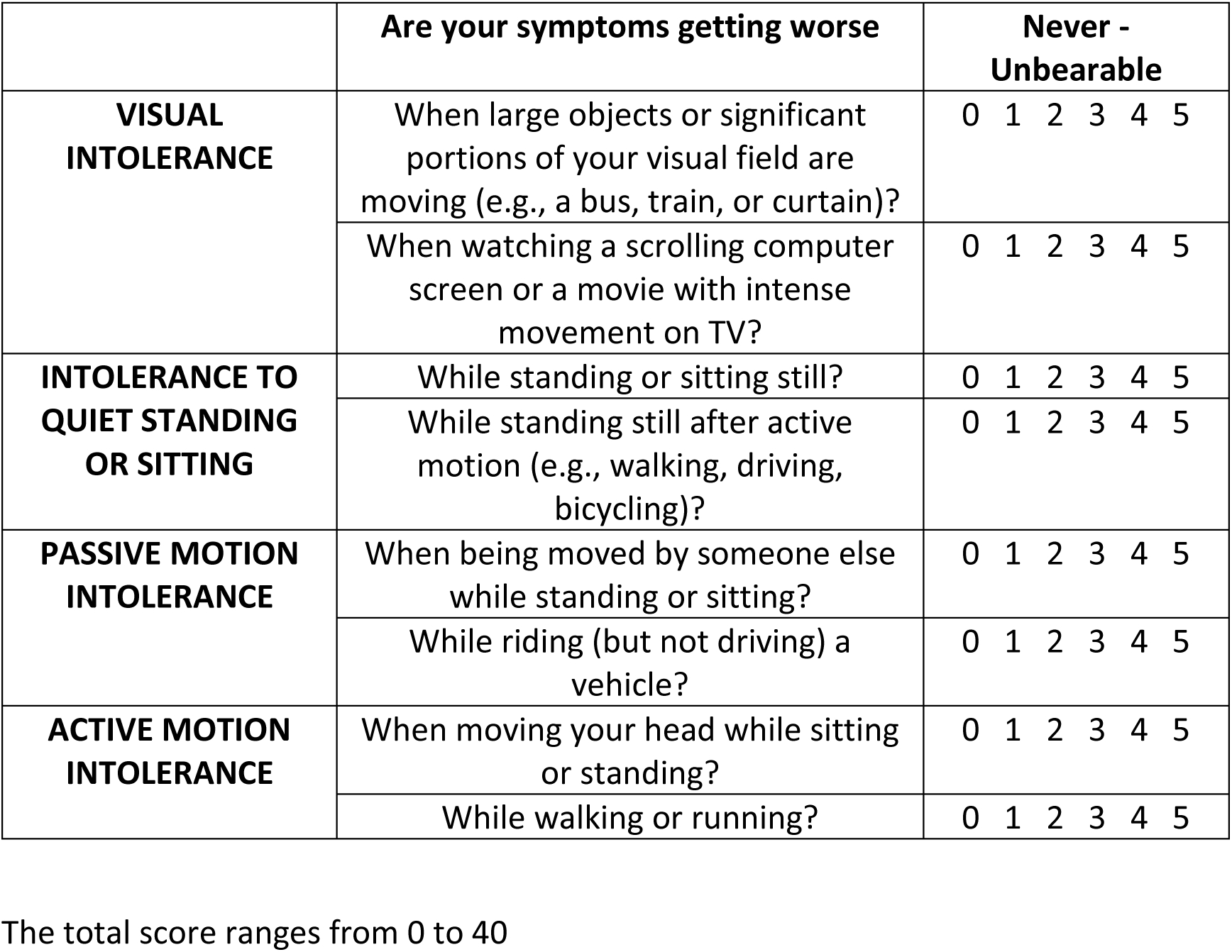
ALQ (Athens-Lübeck Questionnaire) for subtyping PPPD. Please mark the number that best reflects how much you have been bothered during the past week in the following questions

|  | **Are your symptoms getting worse** | **Never -**<br>**Unbearable** |
| --- | --- | --- |
| **VISUAL**<br>**INTOLERANCE** | When large objects or significant portions of your visual field are moving (e.g., a bus, train, or curtain)? | 0 1 2 3 4 5 |
|  | When watching a scrolling computer screen or a movie with intense movement on TV? | 0 1 2 3 4 5 |
| **INTOLERANCE TO**<br>**QUIET STANDING**<br>**OR SITTING** | While standing or sitting still? | 0 1 2 3 4 5 |
|  | While standing still after active motion (e.g., walking, driving, bicycling)? | 0 1 2 3 4 5 |
| **PASSIVE MOTION**<br>**INTOLERANCE** | When being moved by someone else while standing or sitting? | 0 1 2 3 4 5 |
|  | While riding (but not driving) a vehicle? | 0 1 2 3 4 5 |
| **ACTIVE MOTION**<br>**INTOLERANCE** | When moving your head while sitting or standing? | 0 1 2 3 4 5 |
|  | While walking or running? | 0 1 2 3 4 5 |

The total score ranges from 0 to 40

An experienced physician specializing in vestibular medicine explained the procedure to the patients during their visit to the outpatient clinic. Following this, patients completed the questionnaire in a quiet room. They were encouraged to ask any clarifying questions and address potential misunderstandings during or after completing the questionnaire and were allowed to revise their answers if needed.

Descriptive analysis was employed to summarize the distribution of responses for each item. Cronbach’s alpha was used as a reliability coefficient to measure the internal consistency of the ALQ. Interrelations among the different items were analyzed by constructing correlation matrices. After extracting the four symptom subtypes from the eight ALQ items, we assessed the presence of dominant subtypes and determined the frequency of each subtype. A repeated-measures ANOVA was conducted to assess differences in the average symptom severity across the different PPPD subtypes. Chi-square tests were used to evaluate differences in the frequency of occurrence of each symptom subtype. To evaluate how effectively the four proposed subtypes classify our PPPD cohort into these four categories, we conducted a discriminant analysis. In a subgroup of patients, we also utilized the NPQ. For this group, we examined the relationship between the total score and subitem scores of both the NPQ and the ALQ using Pearson correlation analysis. Statistical analyses were performed using SPSS software version 21.0 (IBM, Armonk, New York). Statistical differences were considered significant for p-values less than 0.05. Data visualization was carried out using SPSS, SRplot [22], and the STHDA platform (http://www.sthda.com/english/).

## Results

A total of 96 patients participated (64 female, 32 male), with a mean age of 50.3 years (range 22– 87). Of these, 41 had primary PPPD, while 55 had secondary PPPD, the latter being a consequence of a preceding vestibular disorder. The mean duration of PPPD was 45.5 months (range 3–300 months).

All participants completed the ALQ in full (ALQ 1-8, Table 1). With a theoretical maximum score of 40, the mean total ALQ score was 14.8 (standard deviation 8.2). The internal consistency of the questionnaire, as measured by Cronbach’s alpha, was considered good (α = 0.813), indicating the reliability of the selected items. Detailed percentages for each stage of the 1–5 Likert scale, for each item separately, are illustrated in Figure 1. We further examined the relationship between each questionnaire item by calculating an 8 x 8 correlation matrix. As shown in Figure 2, strong correlations were primarily found between item pairs within the same symptom subtype (e.g., ALQ1 and ALQ2, ALQ3 and ALQ4, ALQ5 and ALQ6, ALQ7 and ALQ8), while weaker correlations were observed between items of different subtypes.

**Figure 1.**
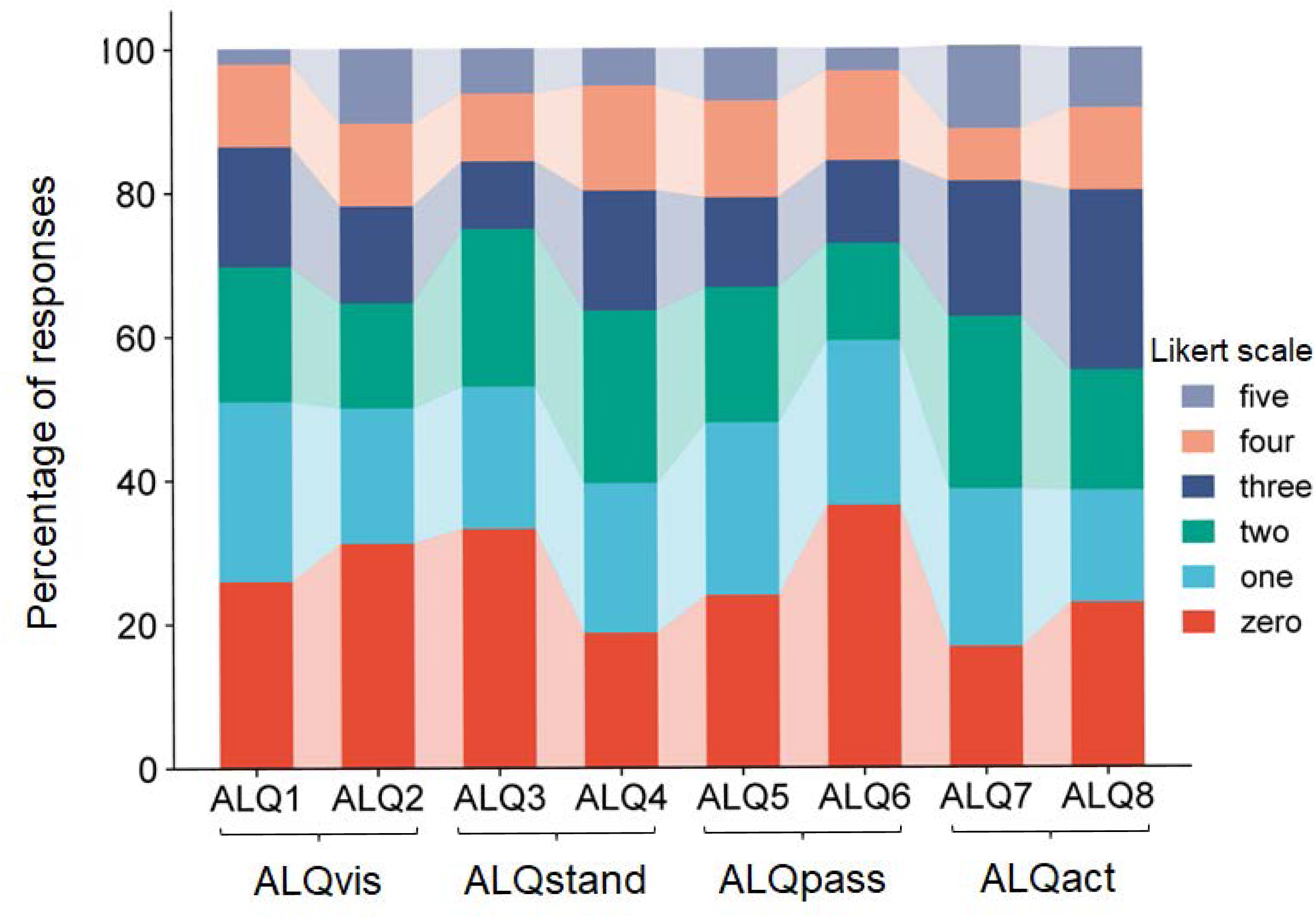
Stacked bar plot showing the percentage occurrence of each Likert scale step for the eight ALQ items (ALQ1 to ALQ8). It can be seen that only a few subjects selected the maximum score, with most preferring lower Likert values.

**Figure 2.**
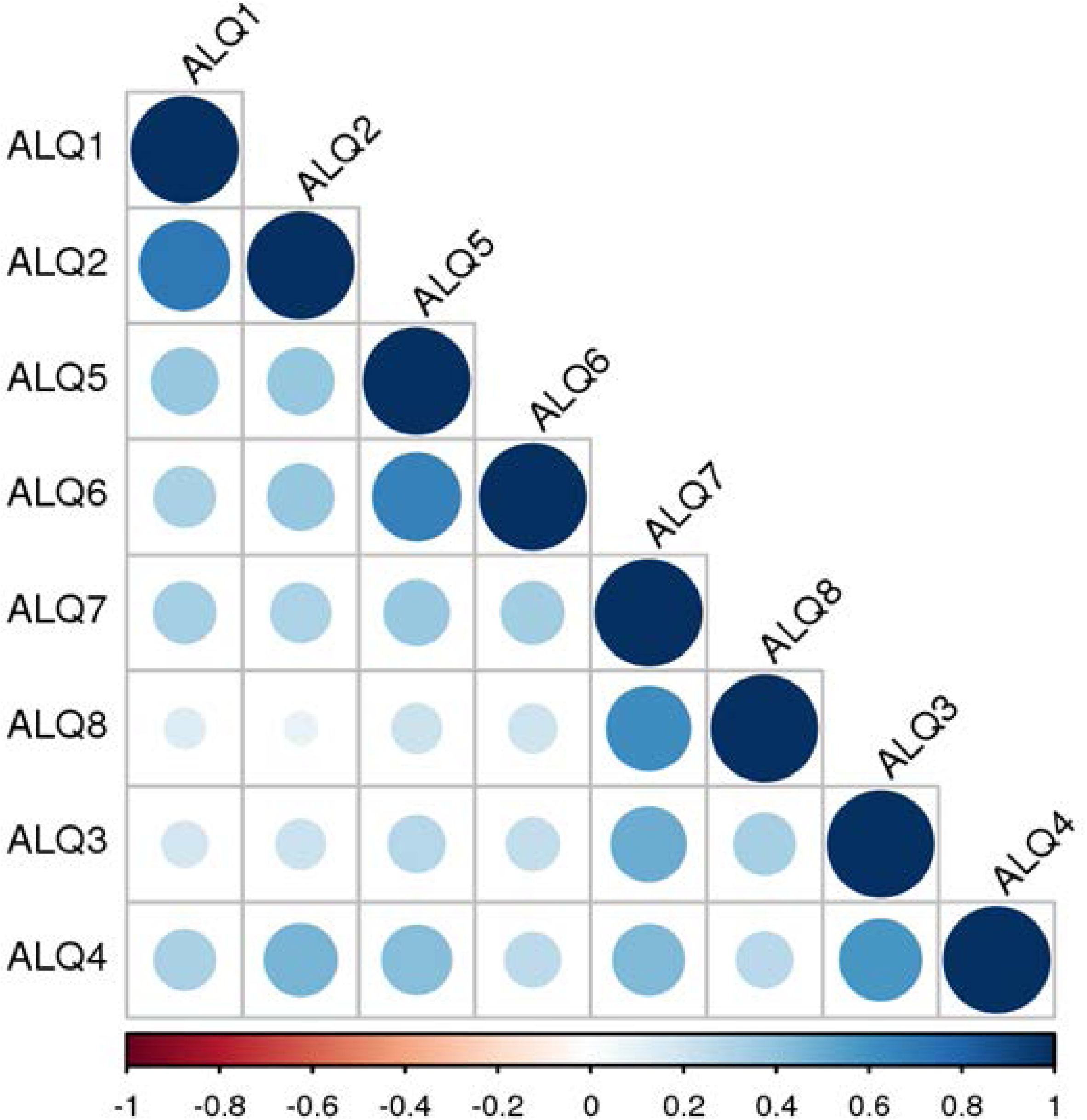
Correlation matrix showing the strength of associations between the eight ALQ items (ALQ1 to ALQ8). Pairs of items belonging to the same symptom domain - ALQ1 and ALQ2 (ALQvis), ALQ3 and ALQ4 (ALQstand), ALQ5 and ALQ6 (ALQpass), and ALQ7 and ALQ8 (ALQact) - exhibit high correlation values. Higher r values are represented by both the diameter of the discs and their color. Positive correlations are shown in shades of blue, ranging from light to dark, while negative correlations (if present) would appear in shades of red, also ranging from light to dark. Notably, no negative correlations were observed.

Active motion intolerance showed, on average, higher scores than the other three symptom domains; however, this difference was only significant on a trend level (F = 2.606, p = 0.052). The mean scores for each symptom subtype - ALQvis, ALQstand, ALQpass, and ALQact - are presented in Table 2. With a theoretical maximum score of 10 for each subtype, some subjects scored zero while others chose the maximum value. Most score distributions, however, were skewed towards lower values, except for ALQact, which exhibited a peak around the mid-range (figure 3A).

**Figure 3.**
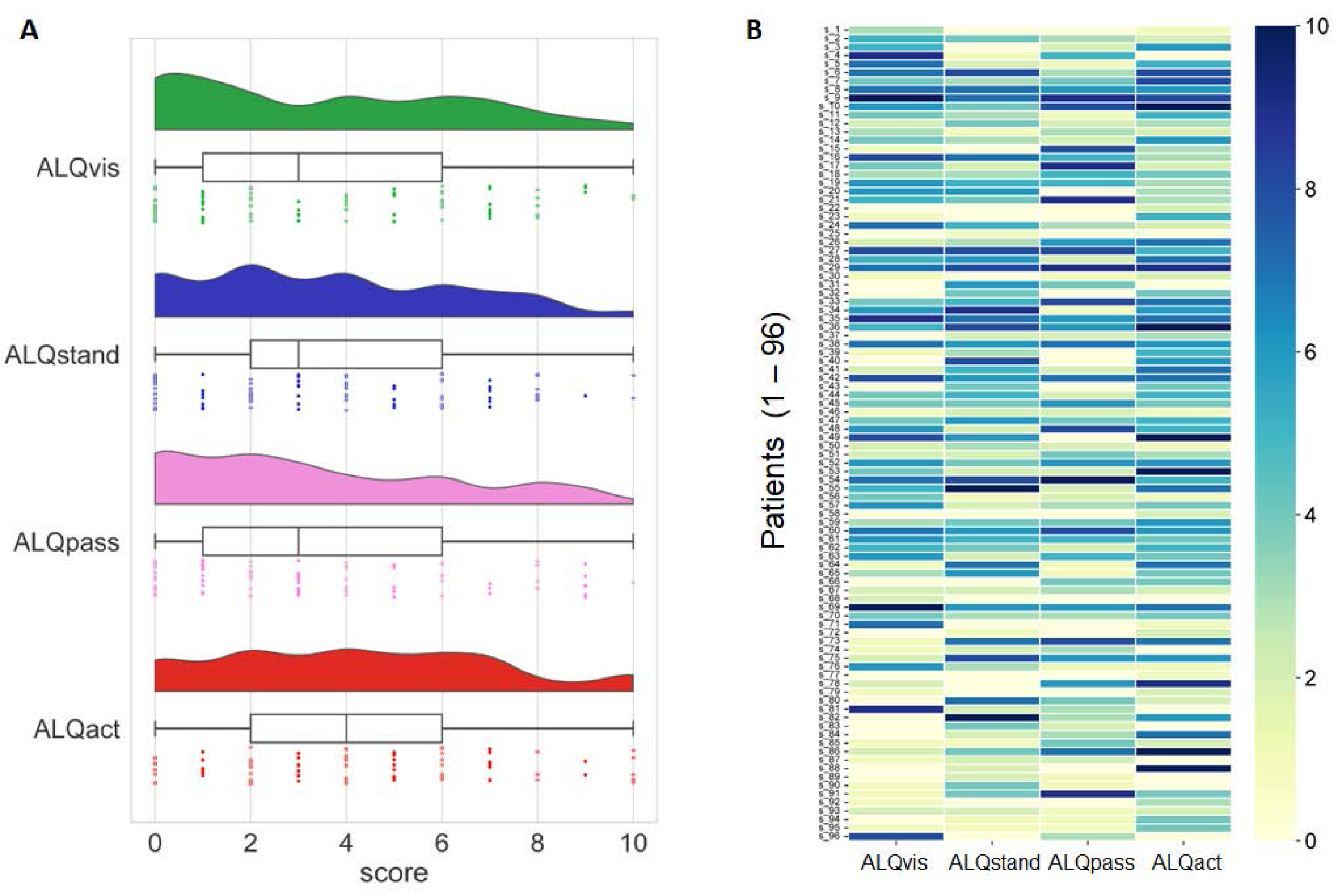
(A) Distribution of ALQ data across the four symptom domains (ALQvis, ALQstand, ALQpass, and ALQact), represented using three methods: kernel density plots, box plots, and individual data points from the 96 subjects. (B) Scores (range: 0 to 10) for each of the four symptom domains (ALQvis, ALQstand, ALQpass, and ALQact) are shown for each of the 96 subjects. Most subjects scored high in only one or, at most, two symptom domains, with lower scores in the remaining domains.

**Table 2.** ALQ (Athens-Lübeck Questionnaire) scores for each symptom subtype— ALQvis (visual intolerance), ALQstand (intolerance to quiet standing or sitting), ALQpass (passive motion intolerance) and ALQact (active motion intolerance).

| | Mean score (± SD) | Range |
| --- | --- | --- |
| **ALQvis** | 3.55 ± 2.92 | 0 - 10 |
| **ALQstand** | 3.64 ± 2.70 | 0 - 10 |
| **ALQpass** | 3.40 ± 2.87 | 0 - 10 |
| **ALQact** | 4.24 ± 2.83 | 0 - 10 |

Examining the scores of each patient individually (Figure 3B) revealed that many participants predominantly fall into one symptom subtype. To identify potentially dominant symptom subtypes from the ALQ, we defined a subject as having a predominant PPPD phenotype only if the relative difference between the highest score and the next highest score was at least 20%. Using this criterion, we found that 65 out of 96 patients exhibited a predominant symptom subtype, with the majority presenting the ALQact phenotype, while the fewest fell into the ALQpass phenotype.

However, in 35% of the cohort, no phenotype predominated over another (figure 4). The tendency for the ALQact phenotype to occur more frequently than the other three was not statistically significant when analyzed for the entire group (χ² = 5.226, p = 0.156).

**Figure 4.**
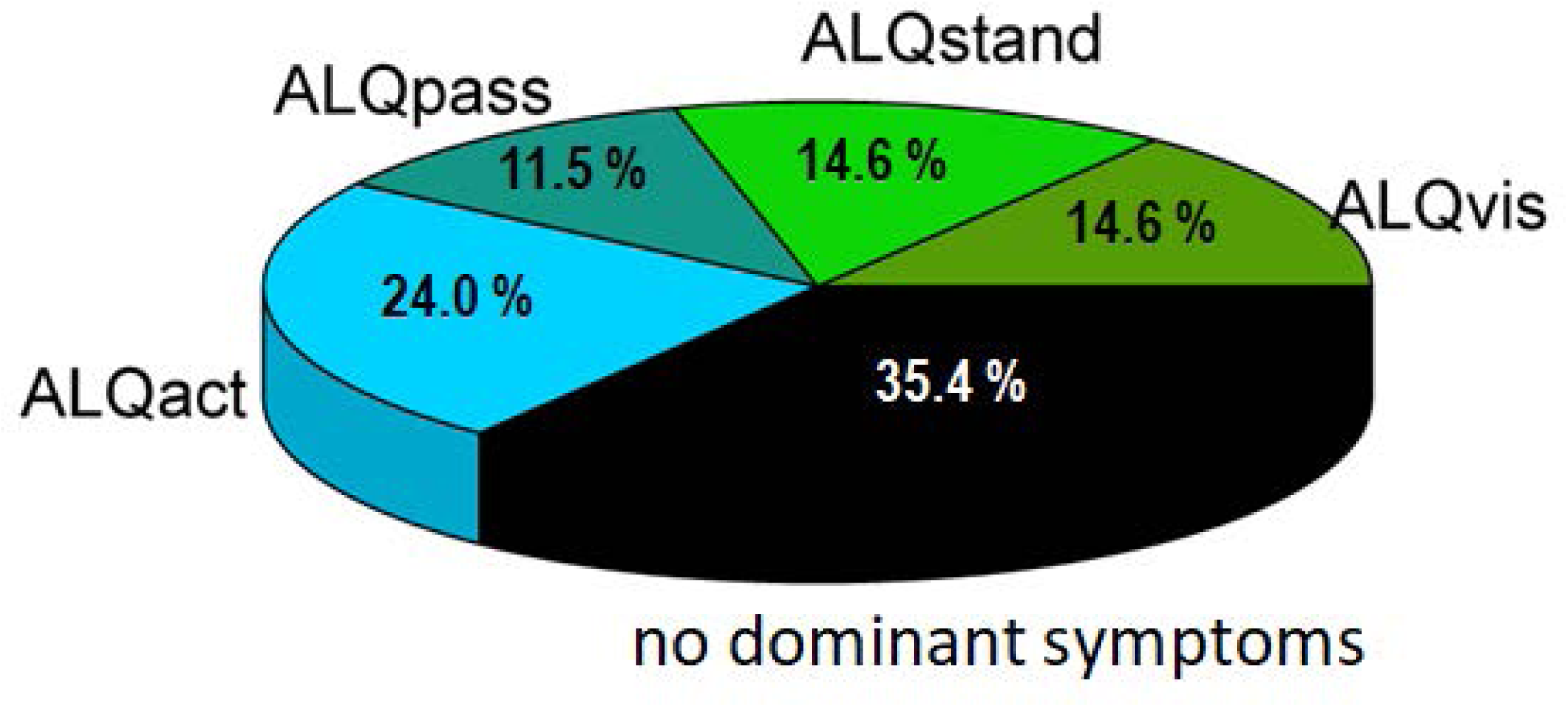
Pie chart showing the distribution of the four symptom subtypes according to the ALQ.

Discriminant analysis demonstrated a clear separation among the different subtypes based on the four question domains (ALQvis, ALQstand, ALQpass, and ALQact), which served as predictors. The analysis yielded significant results (Wilks’ Lambda: χ² =181.6,p <0.001), supporting the existence of the four PPPD subtypes. Table 3 presents the classification function coefficients, and Figure 5 illustrates the canonical discriminant functions. Overall, 77.1% of patients were correctly classified into the predicted subtypes. Specifically, 78.6% were accurately assigned to the ALQvis and ALQstand subtypes, 54.5% were correctly categorized as ALQpass, and 82.6% were accurately included in the ALQact subtype.

**Figure 5.**
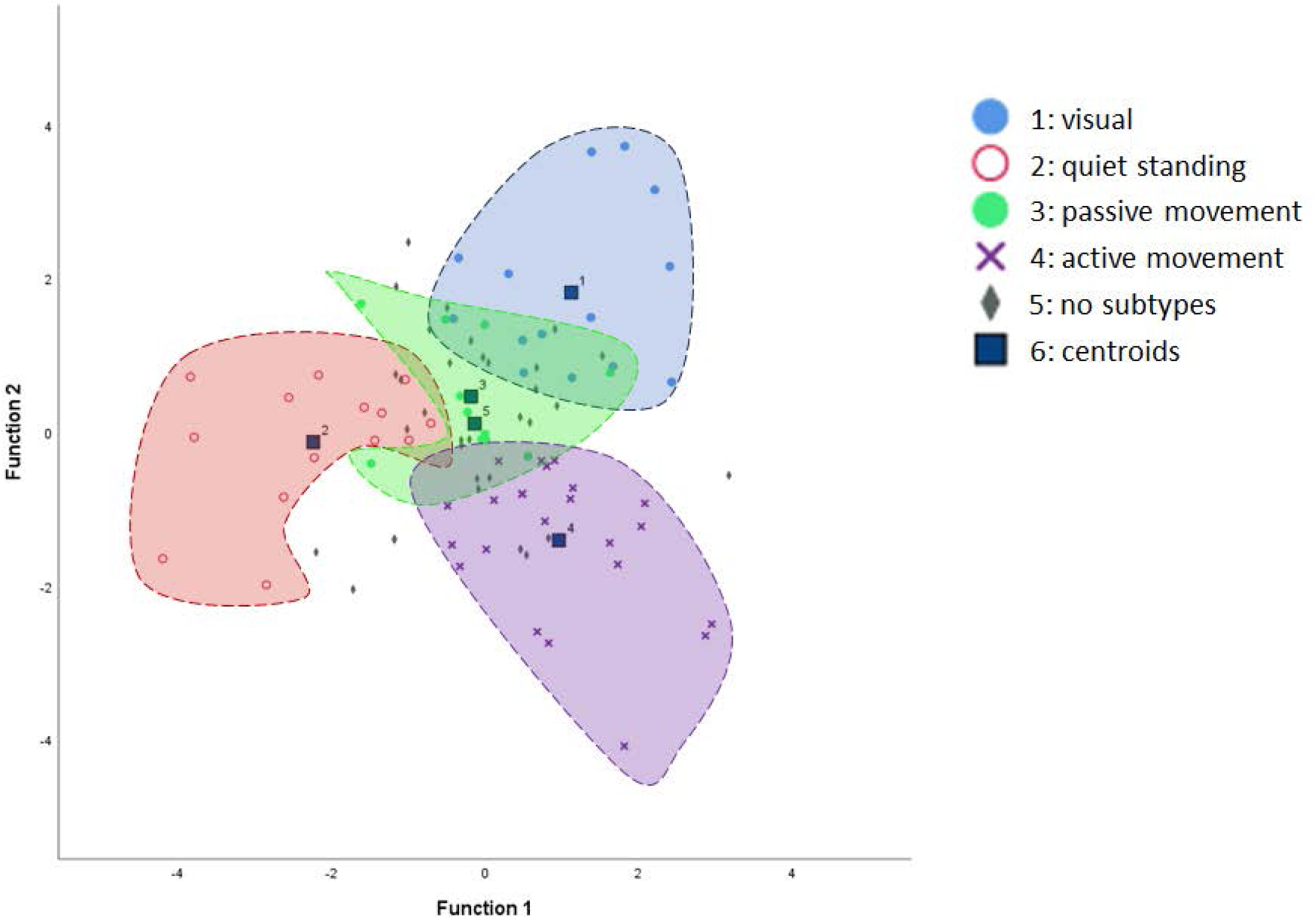
Canonical discriminant function scatter plot illustrating the separation of subtypes. The mean discriminant scores for each subtype are represented as centroids. For clarity, the “no-subtype” category is not enclosed by an inclusion line, as its individual case dots are sparsely intermingled with the rest of the data, which could otherwise obscure the graph.

**Table 3.**
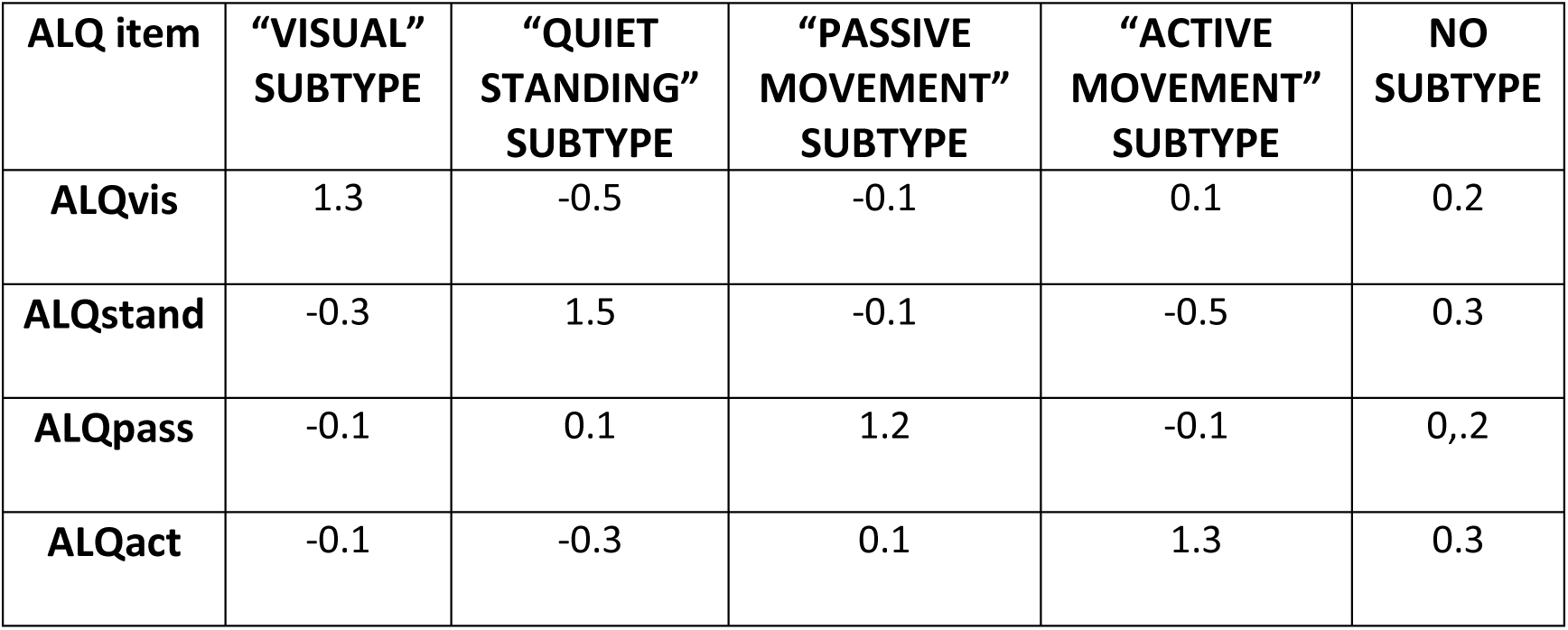
Classification function coefficients based on Fisher’s linear discriminant functions. Each of the four ALQ items shows the highest coefficient for its corresponding PPPD subtype. In contrast, the ALQ items exhibit low coefficients for the “no-subtype” category.

The NPQ was administered to 41 of the 96 patients. Correlation analysis revealed a strong positive linear relationship between the total scores of the ALQ and NPQ (r = 0.834, P < 0.001). Regarding subtype scores, the NPQ “Visual Stimulation” subtype was significantly correlated with ALQvis (r = 0.754, P < 0.001), ALQstand (r = 0.567, P < 0.001), ALQpass (r = 0.598, P < 0.001), and ALQact (r = 0.456, P < 0.01). The NPQ “Movement” subtype showed significant correlations with ALQvis (r = 0.695, P < 0.001), ALQstand (r = 0.435, P < 0.01), ALQpass (r = 0.667, P < 0.001), and ALQact (r = 0.502, P < 0.001). Finally, the NPQ “Upright Posture/Walking” subtype was significantly correlated with ALQstand (r = 0.530, P < 0.001) and ALQact (r = 0.524, P < 0.001) but not with ALQvis (r = 0.194, P > 0.05) or ALQpass (r = 0.192, P > 0.05).

## Discussion

After developing the ALQ to characterize different PPPD symptom subtypes, we demonstrated that it could be easily applied to all patients, without complaints regarding effort or comprehension. The ALQ proved to be internally consistent, supporting the validity of the selected questionnaire items.

Items within the same symptom domain showed higher correlations with each other, as opposed to items belonging to different subtypes. Discriminant analysis, using the four putative phenotypes as predictors, demonstrated a clear classification into the presumed four PPPD subtypes.

Approximately two-thirds of the participants displayed a predominant symptom subtype, with the majority falling into the ALQact phenotype. This indicates that worsening of dizziness with active self-motion of the head or body is particularly prevalent among PPPD patients. The other three subtypes (ALQvis, ALQstand, and ALQpass) were less frequent and tended to be less severe, as participants generally rated their symptom severity lower on the Likert scale. Nevertheless, about one-third of the PPPD cohort could not be classified as having a predominant symptom subtype. Possible reasons include: (i) relatively equal scoring across all four symptom domains, (ii) near-zero responses to all ALQ items, indicating they were not severely affected enough to exhibit a dominant type, or (iii) the existence of additional subtypes with undetermined symptom constellations that were not addressed by our four defined subtypes.

It is also relevant to examine the relationship between the results of the ALQ and the NPQ. While the NPQ was originally designed to assess the severity of PPPD [20], it also indirectly addresses certain putative PPPD subtypes [21]. Our analysis revealed a strong relationship between the total scores of the ALQ and NPQ, indicating a high degree of convergent validity. When examining the correlations between the identified PPPD subtypes, the relationships between individual ALQ and NPQ items proved more challenging to interpret. The most distinct correlation was observed between ALQvis and the NPQ “visual stimulation” subtype. The other ALQ subtypes also showed correlations with most NPQ subtypes, albeit in a less specific manner. This can be attributed to several factors: the NPQ does not address all ALQ subtypes (e.g., it does not differentiate between passive and active motion subtypes), and even where similar subtypes are ostensibly assessed, the NPQ uses a different number of questions and distinct phrasing. These differences likely introduce important confounders that complicate direct comparisons.

The definition of PPPD was a significant collaborative effort among vertigo specialists, with one of its major achievements being the consolidation of the entire spectrum of functional dizziness under a single diagnostic umbrella. This was intended to aid diagnosis, enhance communication of the syndrome to patients, and facilitate planning for multicenter therapeutic studies. Consequently, the question arises as to why we attempted to re-fragment PPPD into symptom subtypes using the ALQ.

The first reason comes from research into the pathophysiological underpinnings of PPPD. Several functional and structural brain imaging studies have implicated different cortical regions in PPPD pathophysiology [23-25], though these studies often show some discrepancies. One reason for this might be the selection of PPPD patients, as it is plausible that, for example, a patient with a predominant visual intolerance subtype would exhibit different dysfunctional brain areas compared to a patient who primarily experiences active motion intolerance. Therefore, incorporating ALQ- based phenotyping into experimental brain imaging studies may help resolve some of these inconsistencies. Examining the PPPD subtypes based on individual patient ratings may also provide insights into the intriguing chicken-and-egg question of whether potential functional or structural brain abnormalities contribute to, or merely reflect, abnormal postural perceptions in PPPD.

A second, and perhaps more significant, reason for classifying PPPD into different symptom subtypes is to facilitate the planning of therapeutic interventions. Since PPPD is not always accompanied by anxiety disorders or depression, it is well-known among vertigo specialists that the use of anti-anxiety or antidepressant medications yields inconsistent results, despite some encouraging findings in smaller studies [26, 27]. Similarly unpredictable is the effectiveness of psychotherapeutic interventions, including cognitive behavioral therapy and patient education, which show benefits in some patients but sometimes with limited long-term efficacy [28, 29]. A multimodal approach, combining medication, psychotherapeutic interventions, and vestibular rehabilitation training, appears to hold the most promise for long-term management of PPPD [30].

Vestibular rehabilitation, a cornerstone of PPPD treatment, encompasses a variety of vestibular exercises designed to recalibrate the dysfunctional equilibrium typical in PPPD. Inspired by the seminal work of Cawthorne and Cooksey [31], modern physiotherapeutic interventions for vestibular rehabilitation have become vast and varied, both in physical and virtual reality settings [32, 33]. These include exercises to promote gaze stability (gaze stabilization exercises, sometimes involving adaptation and substitution exercises), exercises to habituate specific symptoms (habituation exercises, including optokinetic exercises), exercises to improve balance, postural reflexes, and gait, and longer walking exercises for endurance. To date, few studies have targeted homogeneous PPPD patient groups, and their results are challenging to compare due to differences in training approaches [30, 34, 35]. It would be reasonable to assume that customizing a tailored exercise program for different PPPD subtypes might enhance therapeutic efficacy and potentially reduce the daily time required for each exercise.

This study has some potential limitations. First, the relatively high proportion (i.e., 35%) of patients without a symptom predominance may indicate either that these patients genuinely have a mixed phenotype with complaints distributed evenly across the four symptom subtypes, or that there are other, unaddressed symptom subtypes not captured by the ALQ. More experience with thorough clinical interviewing and open-ended questioning in the coming years will help gain deeper insights into the subjective dizziness symptoms of PPPD patients. Second, the threshold used to determine predominant subtypes was inevitably arbitrary. We chose a “20% higher than any other subtype” threshold as a reasonable limit after reviewing the entire dataset and observing minimal change in classification between thresholds of 15% to 30%. Depending on the clinical question or planned intervention, other thresholds could be applied to either highlight more nuanced phenotypes with a mix of symptoms or, conversely, to isolate subtypes with very distinct and intense symptoms within a single phenotype.

## Conclusion

The 8-item ALQ is a valid and reliable questionnaire that enables the identification of distinct symptom phenotypes within the PPPD spectrum. Its primary strengths are its brevity and ease of use in an outpatient vertigo clinic setting, allowing for the differentiation of predominant PPPD phenotypes that could be relevant for tailoring future therapeutic interventions.

## Data Availability

All data produced in the present study are available upon reasonable request to the authors.

## Acknowledgements

The study was supported by a research grant from the Deutsche Forschungsgemeinschaft to C.H. (HE 2689/6-1).

## References

1. Staab JP, Eckhardt-Henn A, Horii A, et al. Diagnostic Criteria for Persistent Postural-Perceptual Dizziness (PPPD): consensus document of the committee for the classification of vestibular disorders of the Barany Society. J Vestib Res 2017;27:191 – 208.

2. Brandt T, Dieterich M. Phobischer Attacken Schwankschwindel, ein neues Syndrom? Munch Med Wschr 1986;28:247 – 50.

3. Brandt T. Phobic postural vertigo. Neurology 1996;46:1515–9.

4. Jacob RG, Lilienfeld SO, Furman JMR, Durrant JD, Turner SM. Panic disorder with vestibular dysfunction: further clinical observation and description of space and motion phobic stimuli. J Anxiety Disord 1989;3:117–30.

5. Bronstein AM. Visual vertigo syndrome: clinical and posturography findings. J Neurol Neurosurg Psychiatry 1995;59:472–6.

6. Cousins S, Cutfield NJ, Kaski D, et al. Visual dependency and dizziness after vestibular neuritis. PLoS One 2014;9:e105426.

7. Staab JP, Ruckenstein MJ, Amsterdam JD. A prospective trial of sertraline for chronic subjective dizziness. Laryngoscope 2004;114:1637–41.

8. Staab JP, Ruckenstein MJ. Expanding the differential diagnosis of dizziness. Arch Otolaryngol Head Neck Surg 2007;133:170–6.

9. Söhsten E, Bittar RS, Staab JP. Posturographic profile of patients with persistent postural-perceptual dizziness on the sensory organization test. J Vestib Res 2016;26:319 – 26.

10. Wurthmann, S.; Naegel, S.; Schulte Steinberg, B.; Theysohn, N.; Diener, H.C.; Kleinschnitz, C.; Obermann, M.; Holle, D. Cerebral gray matter changes in persistent postural perceptual dizziness. J. Psychosom. Res. 2017;103: 95–101.

11. Woll J, Sprenger A, Helmchen C. Postural control during galvanic vestibular stimulation in patients with persistent perceptual-postural dizziness. J Neurol. 2019 May;266(5):1236–1249.

12. Teggi R, Gatti O, Cangiano J, Fornasari F, Bussi M. Functional head impulse test with and without optokinetic stimulation in subjects with persistent postural perceptual dizziness (PPPD): preliminary report. Otol Neurotol 2020;41:e70–5.

13. Anagnostou E, Stavropoulou G, Zachou A, Kararizou E. Spectral Composition of Body Sway in Persistent Postural-Perceptual Dizziness. Otol Neurotol. 2021;42(9):e1318–e1326.

14. Schröder L, von Werder D, Ramaioli C, Wachtler T, Henningsen P, Glasauer S, Lehnen N. Unstable Gaze in Functional Dizziness: A Contribution to Understanding the Pathophysiology of Functional Disorders. Front Neurosci. 2021 Jul 20;15:685590.

15. Castro P, Bancroft MJ, Arshad Q, Kaski D. Persistent Postural-Perceptual Dizziness (PPPD) from Brain Imaging to Behaviour and Perception. Brain Sci. 2022 Jun 8;12(6):753.

16. Yagi C, Morita Y, Yamagishi T, Ohshima S, Izumi S, Takahashi K, Watanabe M, Itoh K, Suzuki Y, Igarashi H, Horii A. Changes in functional connectivity among vestibulo-visuo-somatosensory and spatial cognitive cortical areas in persistent postural-perceptual dizziness: resting-state fMRI studies before and after visual stimulation. Front Neurol. 2023 Jul 24;14:1215004.

17. Armenis G, Zachou A, Anagnostou E. Slow stepping rate in the Unterberger test in persistent postural-perceptual dizziness. J Neurol 2023;270:555–558.

18. Murofushi T, Goto F, Ushio M. Habituation disorders in auditory middle latency response of persistent postural-perceptual dizziness patients. Front Neurol. 2024 Mar 6;15:1366420.

19. Storm R, Krause J, Blüm SK, Wrobel V, Frings A, Helmchen C, Sprenger A. Visual and vestibular motion perception in persistent postural-perceptual dizziness (PPPD). J Neurol. 2024 Jun;271(6):3227–3238.

20. Yagi C, Morita Y, Kitazawa M, Nonomura Y, Yamagishi T, Ohshima S, Izumi S, Takahashi K, Horii A. A Validated Questionnaire to Assess the Severity of Persistent Postural-Perceptual Dizziness (PPPD): The Niigata PPPD Questionnaire (NPQ). Otol Neurotol. 2019;40:e747–e752.

21. Yagi C, Morita Y, Kitazawa M, Yamagishi T, Ohshima S, Izumi S, Takahashi K, Horii A. Subtypes of Persistent Postural-Perceptual Dizziness. Front Neurol. 2021;12:652366.

22. Tang D, Chen M, Huang X, Zhang G, Zeng L, Zhang G, Wu S, Wang Y. SRplot: A free online platform for data visualization and graphing. PLoS One. 2023 9;18(11):e0294236. doi: 10.1371/journal.pone.0294236.

23. Indovina I, Riccelli R, Chiarella G, Petrolo C, Augimeri A, Giofrè L, Lacquaniti F, Staab JP, Passamonti L (2015) Role of the insula and vestibular system in patients with chronic subjective dizziness: an fMRI study using sound-evoked vestibular stimulation. Front Behav Neurosci 9:334. 10.3389/fnbeh.2015.00334

24. Wurthmann S, Naegel S, Steinberg BS, Theysohn N, Diener HC, Kleinschnitz C, Obermann M (2017) Cerebral gray matter changes in persistent postural perceptual dizziness. J Psychosom Res 103:95–101. 10.1016/j.jpsychores.2017.10.007

25. Nigro S, Indovina I, Riccelli R, Chiarella G, Petrolo C, Lacquaniti F, Staab JP, Passamonti L (2019) Reduced cortical folding in multi-modal vestibular regions in persistent postural perceptual dizziness. Brain Imaging Behav 13:798–809

26. Staab JP, Ruckenstein MJ, Amsterdam JD. A prospective trial of sertraline for chronic subjective dizziness. Laryngoscope 2004;114:1637–1641

27. Horii A, Uno A, Kitahara T, Mitani K, Masumura C, Kizawa K et al. Effects of fluvoxamine on anxiety, depression, and subjective handicaps of chronic dizziness patients with or without neuro-otologic diseases. J Vestib Res 2007;17(1):1–8

28. Mahoney AEJ, Edelman S, Cremer PD. Cognitive behavior therapy for chronic subjective dizziness: longer-term gains and predictors of disability. Am J Otolaryngol 2013;34(2):115–120

29. Suica Z, Behrendt F, Ziller C, Gäumann S, Schädler S, Hilfiker R, Parmar K, Gerth HU, Bonati LH, Schuster-Amft C. Comparative effectiveness of non- pharmacological treatments in patients with persistent postural-perceptual dizziness: a systematic review and effect sizes analyses. Front Neurol. 2024;15:1426566.

30. Axer H, Finn S, Wassermann A, Guntinas-Lichius O, Klingner CM, Witte OW. Multimodal treatment of persistent postural-perceptual dizziness. Brain Behav. 2020;10(12):e01864.

31. Cawthorne T. Vestibular injuries. Proc R Soc Lond B Biol Sci 1946;39(5):270–273.

32. Meldrum D, Murray D, Vance R, Coleman S, McConnell S, Hardiman O, McConn Walsh R. Toward a Digital Health Intervention for Vestibular Rehabilitation: Usability and Subjective Outcomes of a Novel Platform. Front Neurol. 2022;13:836796

33. Hall CD, Herdman SJ, Whitney SL, Anson ER, Carender WJ, Hoppes CW, Cass SP, Christy JB, Cohen HS, Fife TD, Furman JM, Shepard NT, Clendaniel RA, Dishman JD, Goebel JA, Meldrum D, Ryan C, Wallace RL, Woodward NJ. Vestibular Rehabilitation for Peripheral Vestibular Hypofunction: An Updated Clinical Practice Guideline From the Academy of Neurologic Physical Therapy of the American Physical Therapy Association. J Neurol Phys Ther. 2022;46(2):118–177.

34. Nada EH, Ibraheem OA, Hassaan MR. Vestibular Rehabilitation Therapy Outcomes in Patients With Persistent Postural-Perceptual Dizziness. Ann Otol Rhinol Laryngol. 2019;128(4):323–329.

35. Choi SY, Choi JH, Oh EH, Oh SJ, Choi KD. Effect of vestibular exercise and optokinetic stimulation using virtual reality in persistent postural-perceptual dizziness. Sci Rep. 2021;11(1):14437.

